# Integrating Histologic Descriptors into the Ninth Edition TNM Staging Improves Prognostic Stratification of Lung Adenocarcinoma

**DOI:** 10.64898/2026.02.17.26346481

**Authors:** Hanie Abolfathi, Michael Maranda-Robitaille, Fabien Claude Lamaze, Manal Kordahi, Victoria Saavedra Armero, Michèle Orain, Pierre Olivier Fiset, David Joubert, Patrice Desmeules, Andréanne Gagné, Yasushi Yatabe, Yohan Bossé, Philippe Joubert

## Abstract

**Background:** Histologic descriptors such as lymphovascular invasion (LVI), visceral pleural invasion (VPI), spread through air spaces (STAS), and grading system have each been associated with adverse outcomes in lung adenocarcinoma (LUAD). However, with the exception of VPI, these features are not formally incorporated into the TNM staging system. We evaluated the prognostic value and incremental contribution of these histologic descriptors within the framework of the 9^th^ edition TNM staging system.

**Methods:** In total, 1,745 individuals diagnosed with stage I–III invasive non-mucinous lung adenocarcinoma (NM-LUAD) were included in this study, comprising 1139 French-Canadian patients who underwent surgical resection at IUCPQ–Université Laval (discovery cohort) and 606 patients from the National Cancer Center Hospital in Tokyo, Japan (validation cohort). The objective of this study was to assess the prognostic contribution of histologic descriptors, including STAS, and LVI, as complements to conventional 9^th^ edition TNM staging.

**Results:** Grade 3 tumors, LVI, and STAS were identified in 880 (50.4%), 809 (46.4%), and 775 (44.4%) of 1745 cases, respectively. Histologic grade and LVI demonstrated the strongest associations, particularly in early-stage disease, while STAS exhibited a stage-dependent effect, being more impactful in stages II-III. VPI showed less consistent prognostic value. Incorporating these histologic descriptors into TNM staging improved prognostic model performance, with the largest gains driven by histologic grade and LVI, while STAS provided additional, complementary prognostic refinement.

**Conclusion:** These findings demonstrate that key histologic descriptors—including grading system, LVI, and STAS—represent robust and reproducible prognostic parameters. Importantly, these descriptors provide complementary, stage-dependent information that may enhance risk stratification and inform refinement of future TNM staging frameworks, including the forthcoming 10^th^ edition.

## Introduction

Since the development of the staging system by Denoix in 1953 [1], the TNM staging system has undergone three revisions in the past decade, with the 9^th^ edition (2024) introducing updated stage subdivisions and distinctions in metastasis [2, 3]. The 9^th^ edition of the TNM staging for lung cancer represents the most recent step in the ongoing refinement of prognostic frameworks [2, 3].

The International Association for the Study of Lung Cancer (IASLC) is considering the inclusion of non-anatomic prognostic descriptors to improve the prognostic accuracy of the TNM staging system in the upcoming 10^th^ edition. Lymphovascular invasion (LVI), spread through air spaces (STAS), and visceral pleural invasion (VPI) have each shown prognostic relevance in lung adenocarcinoma [4–7].

VPI is currently incorporated into the TNM staging as a T descriptor, and LVI and perineural invasion (Pn) are listed as optional descriptors by the American Joint Committee on Cancer (AJCC) and the Union for International Cancer Control (UICC) [3, 8–11]. Additional histopathologic features, such as STAS, have been proposed for consideration by the IASLC Pathology Committee [12–14]. Several strategies have been explored to integrate these descriptors into TNM staging, including refinement of tumor size cutoffs, modification of T descriptors, and incorporation of histologic risk features as complementary prognostic modifiers [6, 10, 12, 15–18].

Despite recent advancements in the TNM staging system, the incorporation of histology-based features such as grading, LVI, and STAS as formal staging descriptors remains controversial, and their combined prognostic impact across TNM stages I–III under the 9^th^ edition has not been fully elucidated. Moreover, there is still no consensus on how these features should be incorporated into the TNM framework, especially for resected tumors at more advanced stages. Understanding their combined prognostic impact could refine stage-based risk stratification and enhance the overall accuracy of the TNM staging system.

We hypothesized that key histologic descriptors—including grading, LVI, STAS, and VPI—represent independent prognostic factors influencing overall survival (OS) and recurrence-free survival in patients with stage I–III LUAD, as defined by the 9^th^ edition of the TNM staging system. Accordingly, the objectives of this study were to evaluate the prognostic value of these descriptors across TNM stages and to assess whether their integration, individually or in combination, improves survival prediction beyond conventional TNM staging alone. Importantly, our goal was not to position any single feature as dominant, but rather to define their relative and complementary contributions within an integrated staging framework and to determine whether selected histologic descriptors may serve as independent prognostic indicators alongside established TNM components such as VPI.

## Materials and Methods

### Patients Cohort Analysis

This study included two independent discovery and validation cohorts of patients diagnosed with LUAD who underwent complete surgical resection (R0). The discovery cohort consisted of 1603 French-Canadian patients who underwent surgical resection at the Institut Universitaire de Cardiologie et De Pneumologie de Québec-Université Laval (IUCPQ-UL) between 2006 and 2021 and part of the LORD cohort. The validation cohort included 782 patients with LUAD treated at the National Cancer Center Hospital (NCCH) in Tokyo, Japan, all of whom underwent curative surgical resection between 2016 and 2018.

Patients were eligible if they met the following criteria: (1) histologically confirmed LUAD, (2) curative (R0) surgical resection with negative margins, and (3) availability of Hematoxylin and Eosin (H&E) stained slides along with adequate tissue for histologic evaluation. Patients were excluded if they met any of the following criteria: (1) stage IV tumor at diagnosis, (2) presence of specific histologic variants of LUAD (including fetal, colloidal, enteric, and mucinous subtypes), (3) neoadjuvant therapy, (4) multifocal or synchronous tumors, or (5) combined carcinoma histology. After applying the predefined inclusion and exclusion criteria to the **discovery cohort**, 464 patients were excluded, leaving 1139 invasive non-mucinous lung adenocarcinoma (NM-LUAD) cases for analysis. Applying the same criteria to the **validation cohort** resulted in 606 invasive NM-LUAD cases being retained for analysis.

The same histopathological review protocols and classification systems were applied to ensure consistency and comparability across both populations. For all patients, demographic data, Overall survival (OS), Recurrence-free survival (RFS), and histopathological features were collected. OS and RFS were selected as primary clinical endpoints. OS was measured from the date of surgery to death from any cause, with follow-up censored at 5 years. RFS was calculated from surgical resection to either tumor recurrence or death, with censoring at 3 years.

**For the discovery cohort**, the study was approved by the ethics committee of the IUCQP-UL (ethics approval number: MP-10-2022-3752, 22156). **For the validation cohort**, the medical research ethics committee of the National Cancer Center approved this study as part of a larger research project (IRB approval number 2010-077). In both cohorts, the institutional committee waived the need for informed consent due to the retrospective design of this study, which involved reviewing patient records and manually examining radiologic and pathological data.

### Histopathological Evaluation

Hematoxylin and eosin (H&E) stained sections from each case were collected and independently evaluated by thoracic pathologists (PJ, YY, PD, AG, POF, MK). Tumors were classified according to the 2021 grading framework proposed by the IASLC for NM-LUAD [19, 20].

The histological assessment included documentation of architectural growth patterns, STAS, LVI, VPI, tumor dimensions, and pathological stage as defined by the AJCC Cancer Staging Manual [2, 12]. Histologic grading was based on the assessment of six architectural growth patterns (lepidic, acinar, papillary, micropapillary, solid, and complex glandular patterns, including cribriform and fused glands). The relative proportion of each pattern was estimated in 5% increments, with the total summing to 100%. Histologic grade was assigned according to the predominant growth pattern and the presence or absence of high-grade patterns, in accordance with IASLC recommendations, classifying tumors as grade 1 (lepidic-predominant), grade 2 (acinar or papillary-predominant without high-grade patterns), or grade 3 (any tumor containing ≥ 20% high-grade patterns, including solid, micropapillary, or complex glandular patterns; including cribriform and fused glands) [19–22]. Tumor dimensions and pathological stage were determined based on the AJCC Cancer Staging Manual (9^th^ edition) [2, 23].

STAS was assessed on routine H&E-stained sections and defined as the presence of micropapillary clusters, solid nests, or single tumor cells within alveolar air spaces beyond the edge of the main tumor, in accordance with IASLC recommendations [8, 12, 23–25]. Care was taken to distinguish true STAS from potential artifacts such as tissue fragmentation or mechanical tumor displacement. STAS was recorded as a binary variable (present vs. absent), as described in previous publications [12, 26].

LVI was also assessed on routine H&E-stained sections according to standardized morphologic criteria. Lymphatic invasion was defined by the presence of tumor cells within the lumen of endothelial-lined lymphatic channels. Vascular invasion was defined by the presence of tumor cells within the lumen and/or infiltrating the wall of venous or arterial vessels, identified by a recognizable endothelial lining and vessel wall structure. Both intratumoral and extratumoral vessels were evaluated, although extratumoral vascular invasion was rare. Care was taken to distinguish true vascular invasion from retraction artifacts. Elastic stains (Verhoeff–Van Gieson) were performed for the assessment of visceral pleural invasion when the tumor was in contact with the pleura and for vascular invasion in selected cases. LVI was recorded as a binary variable (present vs. absent) [23, 27, 28].

VPI was defined by tumor invasion beyond the elastic layer of the visceral pleura (PL1 or PL2), as determined on routine H&E-stained sections, with elastic fiber staining used in selected cases when the elastic layer was not clearly identifiable, in accordance with AJCC and IASLC recommendations [6, 8, 29–31].

### Statistical Analysis

Baseline clinicopathological characteristics were compared between the discovery and validation cohorts. Categorical variables were evaluated using the chi-square test or Fisher’s exact test, as appropriate, while continuous variables were compared using the Kruskal–Wallis test. Comparisons according to histologic invasion patterns, including STAS, LVI, and VPI, were also performed. Continuous variables were analyzed using the Mann–Whitney U test (Wilcoxon rank-sum test), and categorical variables were also assessed using the chi-square test or Fisher’s exact test.

Effect sizes were calculated to provide additional context for statistically significant findings. For categorical variables, Cramer’s V was used in analyses involving contingency tables larger than 2×2, while the Phi coefficient was employed for 2×2 tables. For continuous variables analyzed with the Mann-Whitney U test, the rank-biserial correlation was calculated to estimate the effect size.

Cox proportional hazards regression was used to evaluate survival outcomes, with both univariate and multivariable models applied for OS and RFS. Kaplan-Meier survival curves were generated for patients across pathologic stage I to III tumors, and the log-rank test was used to compare survival outcomes based on whether STAS, LVI, and VPI were present or absent. To evaluate the prognostic performance of the models, we calculated hazard ratios (HRs), concordance index (C-index), and area under the receiver operating characteristic curve (AUC). Prognostic models were constructed by sequentially integrating histologic grade, STAS and LVI into the 9^th^ edition TNM staging system to assess their stepwise and combined contribution to OS and RFS.

Model comparisons were conducted using the *DeLong* test for differences in AUC and a bootstrap-based test for comparisons of C-index values. The *survival* and *survminer* packages were utilized for Cox regression and Kaplan-Meier survival analyses. The *timeROC*, and *pROC* packages were used for the computation and comparison of C-index and AUC values.

Prognostic models were developed in the discovery cohort. Model coefficients derived from the discovery cohort were then fixed and applied to the external validation cohort using a prediction-based approach. Specifically, linear predictors were calculated in the validation cohort using regression coefficients obtained from the discovery cohort, without re-estimation of model parameters. Model performance in the validation cohort was assessed using discrimination metrics, including the C-index and AUC, to evaluate external validity.

All statistical analyses were conducted using two-sided tests, with significance defined as p ≤ 0.05. Computations were carried out in R (version 4.5.0) via RStudio (Boston, MA, USA).

## Results

### Patient Demographics and Outcome

Table 1 summarizes the clinicopathological features of patients included in the discovery and validation cohorts. **The discovery cohort** consisted of 1,139 patients with resected non-metastatic LUAD. The median follow-up duration was 78.1 months, allowing robust assessment of OS and RFS. Based on the 9^th^ edition TNM staging system, most patients had early-stage tumor, with 829 (72.8%) classified as stage I, 208 (18.3%) as stage II, and 102 (8.9%) as stage III.

**Table 1:**
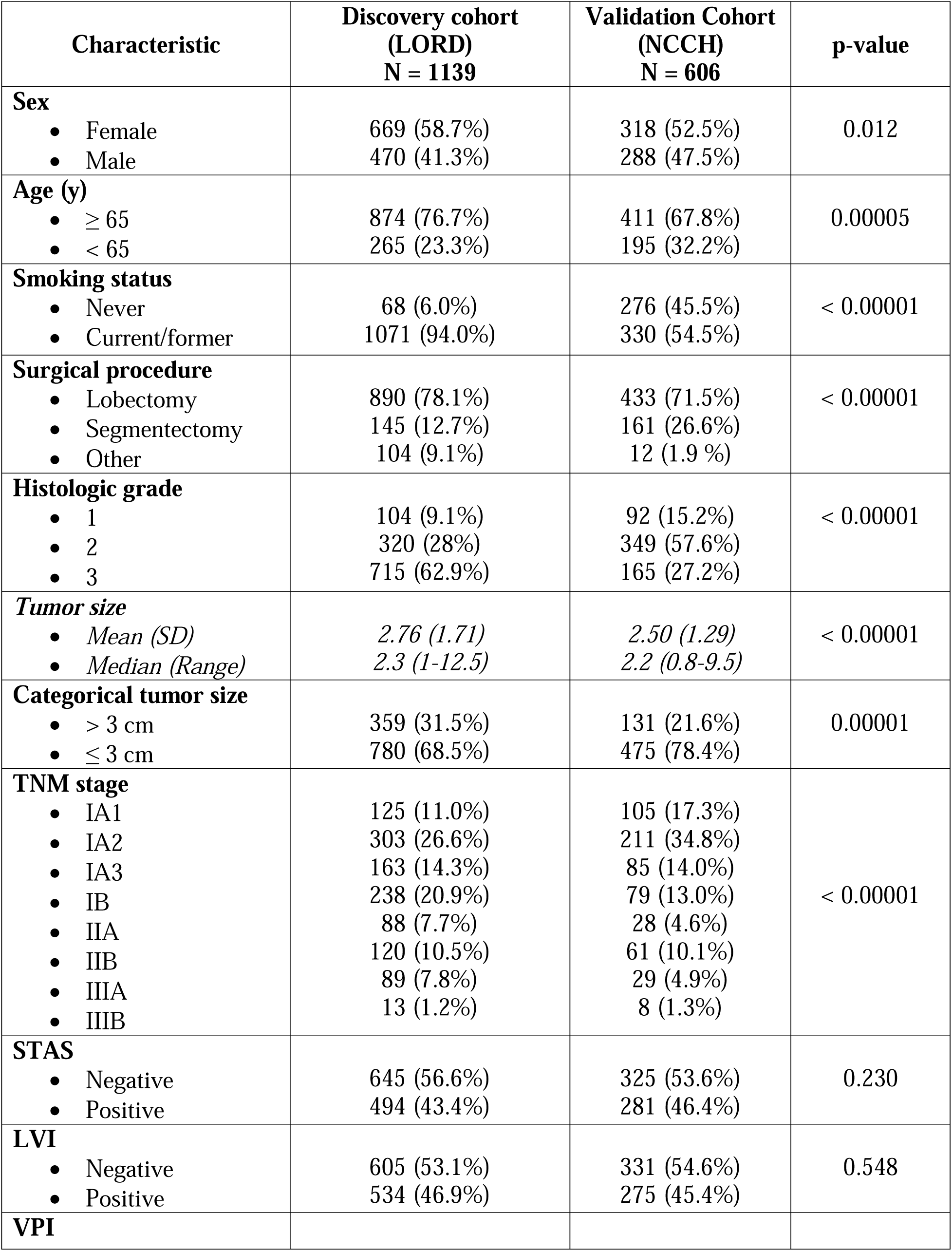
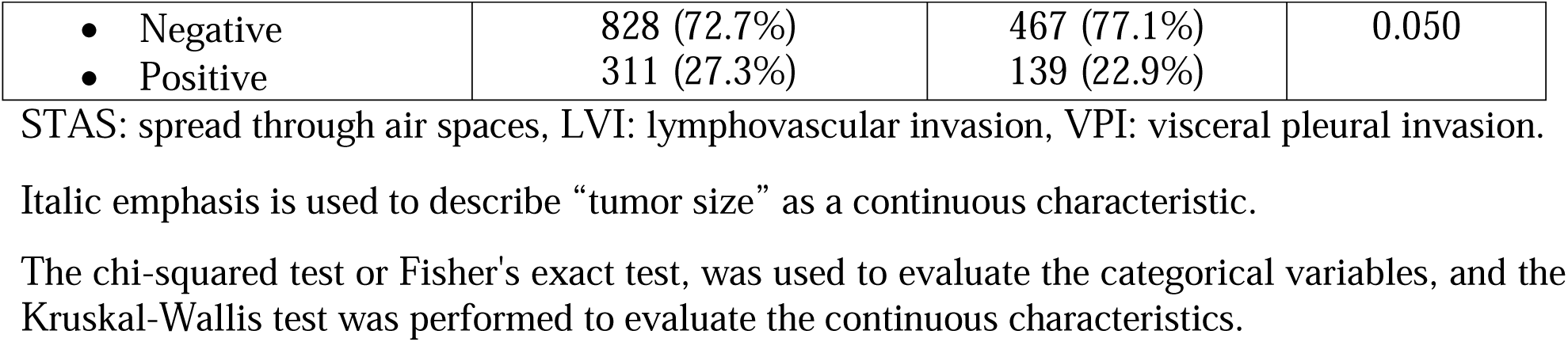
Clinicopathologic Characteristics of Discovery and Validation Cohorts.

Tumor grading according to the 2021 IASLC classification demonstrated that **the discovery cohort** was enriched for high-grade tumors, with 715 cases (62.9%) classified as grade 3, reflecting a substantial representation of aggressive histologic patterns. Tumor size distribution showed that the majority of tumors measured ≤ 3 cm (68.5%), while a smaller proportion exceeded 3 cm (31.5%). At the end of follow-up, 430 patients (37.7%) had died, and 375 (32.9%) had experienced tumor recurrence. Using 5-year censoring, 258 patients (22.6%) had died, while under 3-year censoring for RFS, 217 patients (19.1%) experienced recurrence.

In the **discovery cohort**, STAS (43.4 %) and LVI (46.9 %) were frequently observed, and VPI (27.3 %) was present in a substantial subset of tumors, consistent with an invasive tumor phenotype (**Tables S1 and S2)**. These features were more prevalent among higher-grade tumors and more advanced stages, underscoring their close association with aggressive tumor biology.

To assess the generalizability of our findings, we evaluated a geographically and ethnically distinct **validation cohort**. The prevalence of key invasive histologic descriptors such as STAS, LVI, and VPI were broadly comparable to those of the discovery cohort. **In the validation cohort**, the median tumor size was 2.2 cm (range, 0.8–9.5 cm), and tumors were more frequently ≤ 3 cm (78.4%) than > 3 cm (21.6%). Most patients had stage I tumor (79.2%), with smaller proportions classified as stage II (14.7%) or stage III (6.1%). Compared with the discovery cohort, **the validation cohort** showed a lower prevalence of high-grade tumors, with 27.2% classified as grade 3, as well as a lower prevalence of VPI (22.9%).

### Distribution of STAS, LVI, and VPI Across Clinicopathologic Characteristics

The distribution of STAS, LVI, and VPI across clinicopathologic characteristics in the discovery cohort is summarized in **Table S1**. **In the discovery cohort**, among all examined features, histologic grade showed the strongest association with these invasive descriptors. Higher grades were significantly correlated with increased frequencies of STAS (53.2% in grade 3 vs. 26.9% in grades 1–2; p < 0.00001), LVI (62.1% vs. 21.2%; p < 0.00001), and VPI (34.7% vs. 14.9%; p < 0.00001).

Tumor size and TNM stage also demonstrated consistent positive associations. VPI was more common in tumors larger than 3 cm (38.7%) compared to those 3 cm or smaller (22.1%; p < 0.00001). Similarly, the prevalence of LVI increased with advancing stage, from 37.6% in stage I to 71.6% in stages II–III (p < 0.00001).

Sex and age were also associated with LVI. Males had a higher prevalence of LVI than females (52.1% vs. 43.6%; p = 0.009), and LVI was more common in patients under 65 years of age (52.5% vs. 45.2%; p = 0.038). In addition, smoking status was significantly linked to LVI, with higher rates in current or former smokers (48.0%) compared to never-smokers (29.4%; p = 0.002).

The prevalence of STAS, LVI, and VPI consistently increased with advancing histological grades and TNM stages **(Table S2)**. The overlap and co-occurrence of aggressive histologic features, including high-grade tumors, LVI, STAS, VPI, and advanced TNM stage, are further illustrated in **Figure 1**.

**Figure 1:**
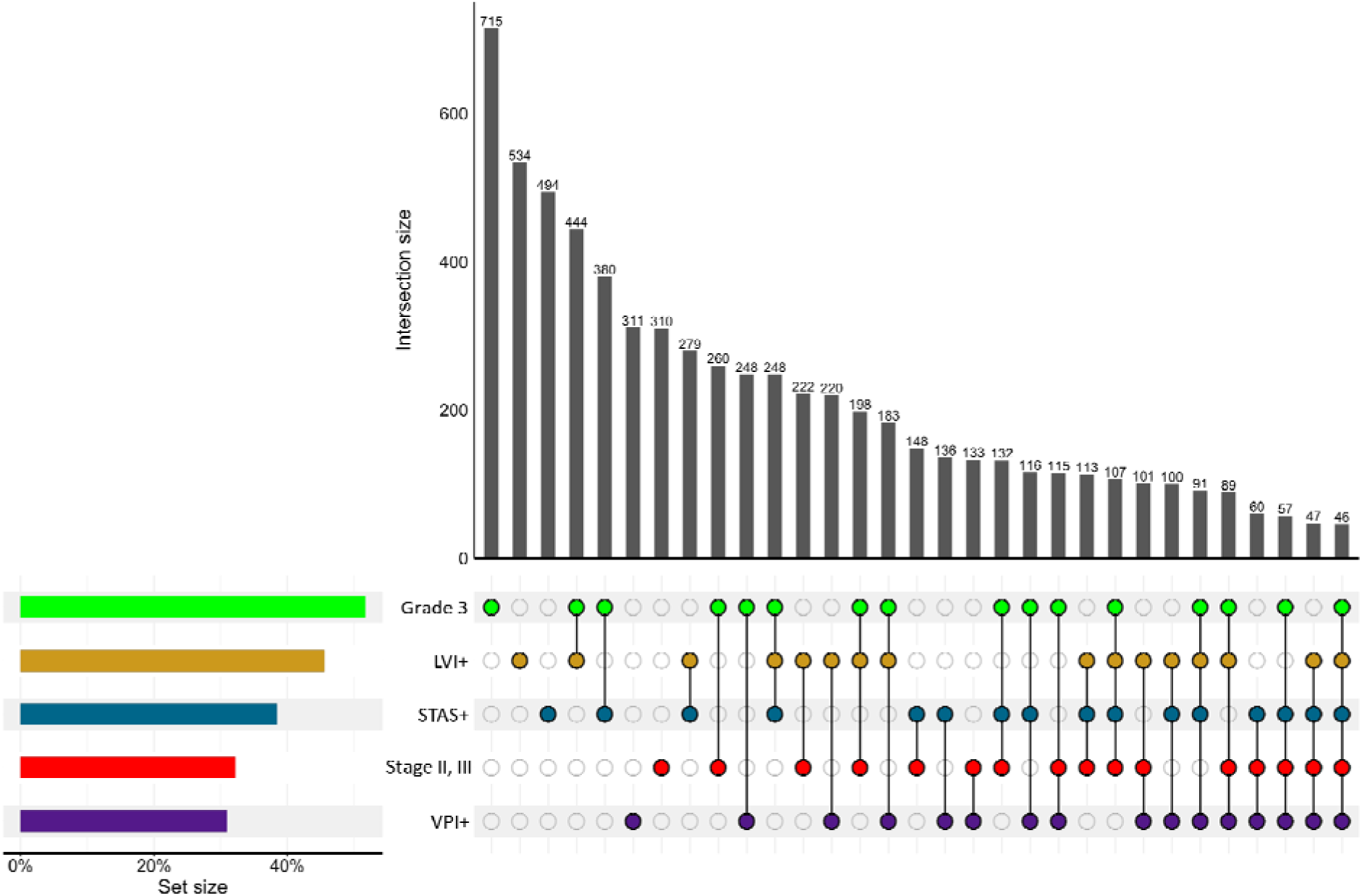
UpSet Plot of Overlapping Histopathological Features in LUAD (LORD cohort, N=1139). The UpSet plot illustrates the intersection of key histopathological features in LUAD patients, including Grade 3 tumors, LVI, STAS, VPI, and tumor stage (II-III). The bar chart at the top represents the intersection size, showing the number of patients exhibiting specific combinations of these features. The connected dots in the lower matrix indicate which features are present in each intersection group. The left-side bar plots display the overall frequency of each individual feature in the dataset. This visualization highlights the co-occurrence of aggressive tumor characteristics and provides insights into their distribution within the patient population. STAS: spread through air spaces, LVI: lymphovascular invasion, VPI: visceral pleural invasion.

Similar results were observed in the **validation cohort**. Histologic grade remained significantly associated with all three histologic descriptors. Specifically, the frequency of STAS, LVI, and VPI increased with higher histologic grade, with statistically significant associations for STAS (p = 0.011), LVI (p = 0.01), and VPI (p = 0.046).

These findings underscore the differential and overlapping relationships of STAS, LVI, and VPI with key clinicopathologic variables, highlighting the need for their collective consideration in lung cancer staging and prognosis.

### Survival Outcomes Based on Histologic grade, STAS, LVI, and VPI

The vital status and date of death were available to calculate OS in all 1139 patients, and a total of 827 of all patients (72.6%) had at least one post-surgery visit for RFS analysis. The median OS for patients positive for STAS, LVI, and VPI was 71.1, 72.0, and 74.6 months, respectively, compared to 82.8, 81.8, and 78.3 months in those without each of these features. Similarly, the median RFS for STAS-, LVI-, and VPI-positive patients was 55.8, 49.0, and 48.8 months, respectively, versus 64.8, 66.6, and 63.8 months respectively in those without each of these features.

Kaplan–Meier survival analyses in the discovery cohort demonstrated that STAS and LVI were each associated with adverse outcomes, although their prognostic effects differed by tumor stage. STAS-positive tumors were associated with significantly worse OS and RFS in the overall population (OS: p = 0.0084; RFS: p = 0.0052; **Figure 2A, B**). Stage-stratified analyses revealed that the adverse impact of STAS on OS was observed in patients with stage II–III tumor (p = 0.0043), with no significant association observed in stage I tumors (p = 0.53). In contrast, STAS was not significantly associated with RFS in either stage I (p = 0.11) or stage II–III tumor (p = 0.10) **(Figure S1)**.

**Figure 2:**
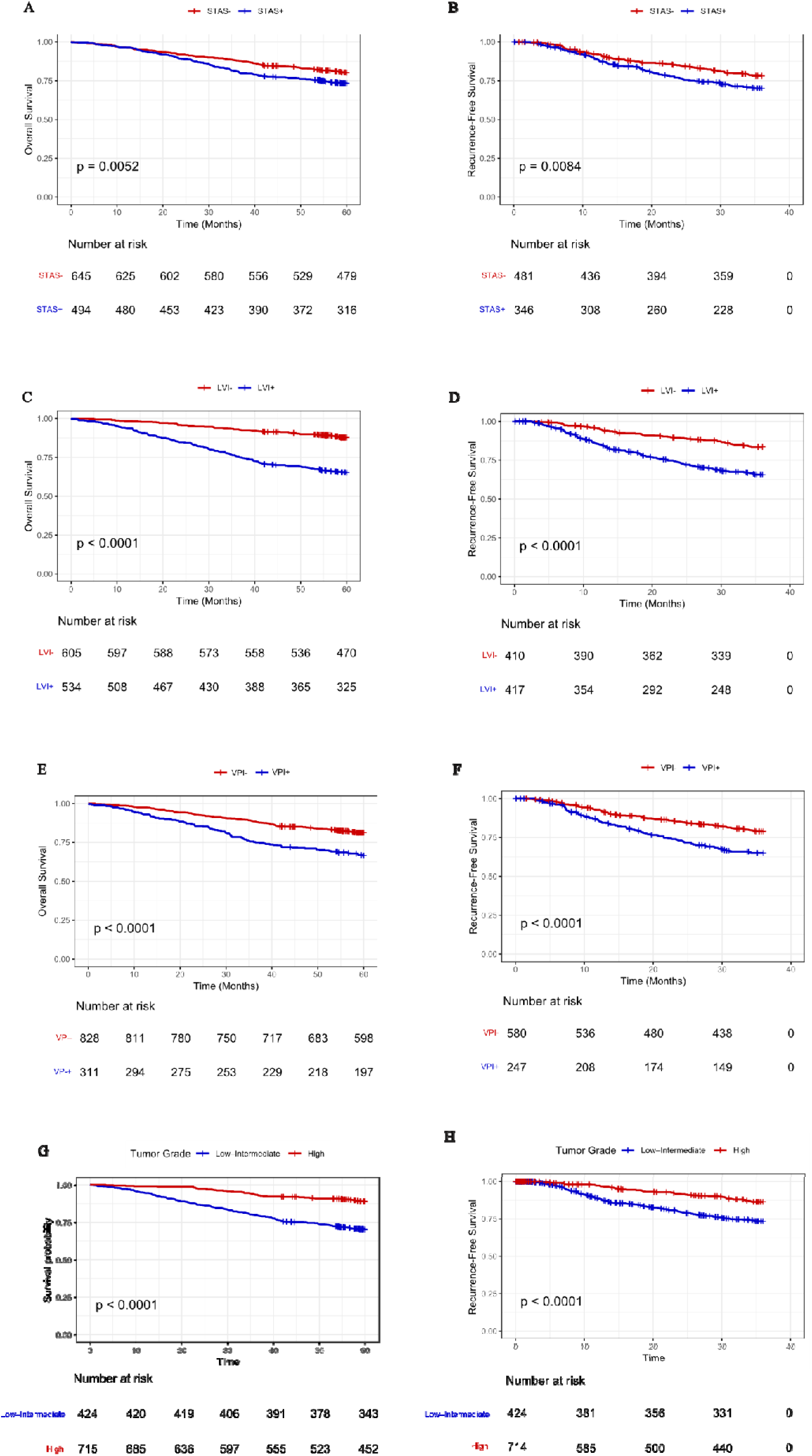
Prognostic significance of STAS, LVI, and VPI for overall survival (OS) and recurrence-free survival (RFS) in patients with resected LUAD from the discovery cohort (N = 1139). **(A, B)** OS and RFS according to STAS status; **(C, D)** OS and RFS according to LVI status; **(E, F)** OS and RFS according to VPI status; **(G, H)** OS and RFS according to the histologic grade. STAS: spread through air spaces, LVI: lymphovascular invasion, VPI: visceral pleural invasion.

LVI was strongly associated with worse OS and RFS in the overall cohort (p < 0.00001 for both; **Figure 2C, D**). Stage-stratified analyses showed that its impact on RFS was most pronounced in stage I tumor (p < 0.0001), whereas no significant association was observed in stages II–III (p = 0.36). In contrast, LVI remained significantly associated with OS across both early and advanced stages (stage I: p < 0.0001; stages II–III: p = 0.00063; **Figure S2**). Together, these findings indicate that STAS and LVI capture complementary aspects of tumor aggressiveness, with stage-dependent prognostic relevance rather than uniform effects across all tumor stages.

For VPI, the VPI negative patients had significantly better OS and RFS compared to those with VPI (p < 0.00001 for both) **(Figure 2. E, F)**. Stratified by stage, VPI was significantly associated with both RFS (p = 0.0023) and OS (p < 0.0001) in stage I, but no significant differences were observed in stages II–III for either RFS (p = 0.42) or OS (p = 0.41) **(Figure S3)**.

Histologic grade was also strongly associated with survival outcomes. Kaplan–Meier analyses demonstrated a stepwise deterioration in both OS and RFS with increasing histologic grade in the overall cohort **(Figure 2G, H)**. Patients with grade 3 tumors exhibited significantly worse OS and RFS compared with those with grade 1–2 tumors (both p < 0.00001).

Stage-stratified analyses revealed that the prognostic impact of histologic grade differed by disease stage **(Figure S4)**. In stage I disease, higher histologic grade was associated with significantly worse OS (p < 0.0001) and RFS (p = 0.00012), indicating that histologic grading captures aggressive tumor biology even in early-stage tumors. In contrast, in stage II–III disease, histologic grade was not significantly associated with either OS or RFS.

These findings indicate that histologic grade, LVI, and STAS provide stage-dependent prognostic information, reflecting distinct biological mechanisms of tumor aggressiveness that dominate at different points in disease progression, rather than uniform effects across all stages.

The multivariable Cox regression model **(Table 2)** supported the results observed in the Kaplan–Meier survival analysis, demonstrating that STAS served as an independent prognostic factor for both RFS (HR = 1.334, 95% CI = 1.004–1.773, p = 0.047) and OS (HR = 1.266, 95% CI = 1.045–1.534, p = 0.015) after adjusting for other clinicopathologic variables. LVI also remained a significant predictor of worse OS (HR = 2.072, 95% CI = 1.594–2.481, p = 1.11×10□□) and RFS (HR = 1.542, 95% CI = 1.111–1.942, p = 0.01), with effect sizes of greater magnitude than those observed for STAS. Moreover, histologic grade emerged as one of the strongest and most consistent independent prognostic factors across both cohorts. Grade 3 tumors were associated with significantly worse OS (HR = 1.626, 95% CI = 1.208-2.087, p = 0.001) and RFS (HR = 1.301, 95% CI = 1.061-1.762, p = 0.008) after adjustment for TNM stage, tumor size, surgical approach, and other histopathologic variables, with effect sizes comparable to or exceeding those observed for STAS and VPI.

**Table 2:**
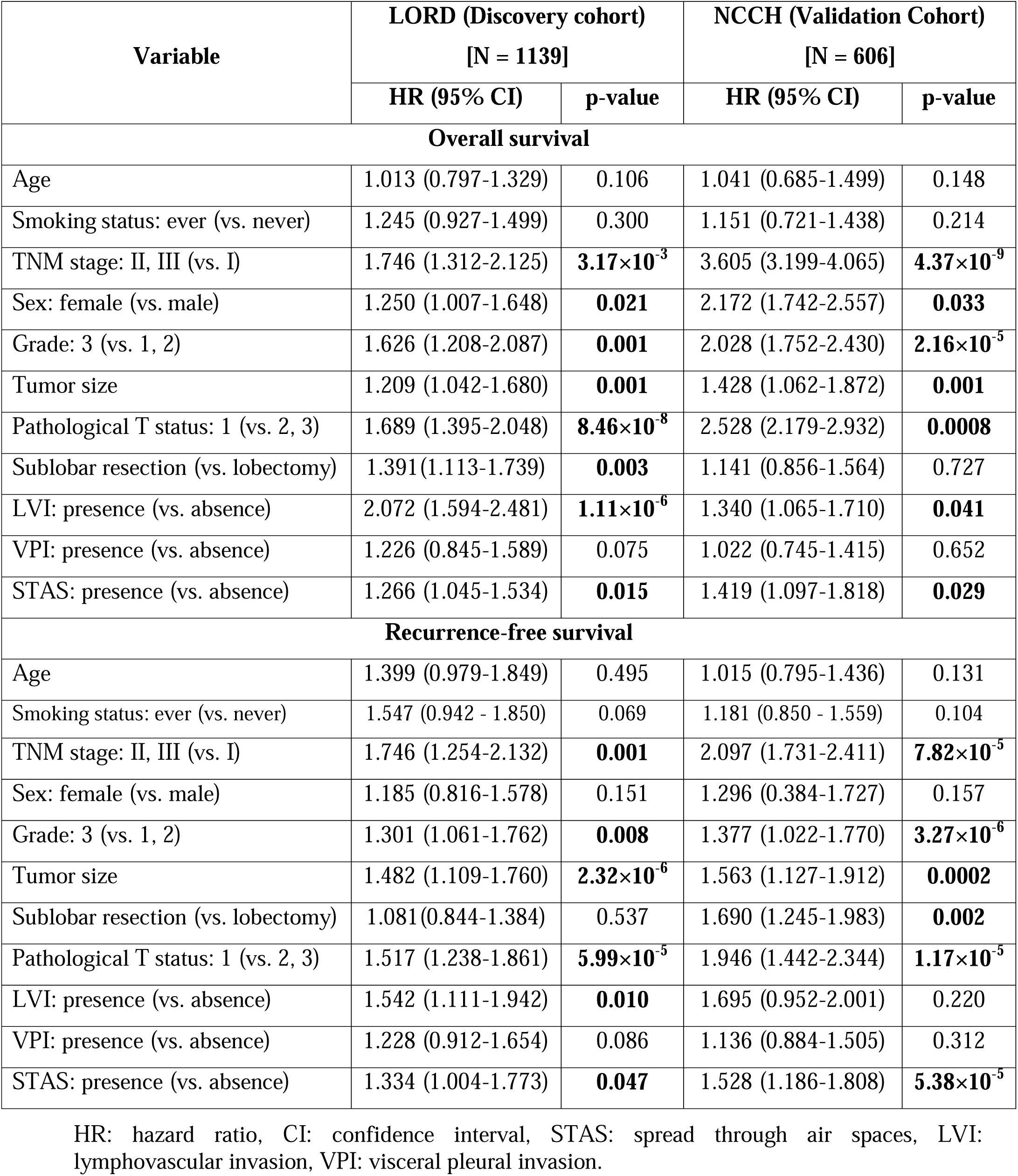
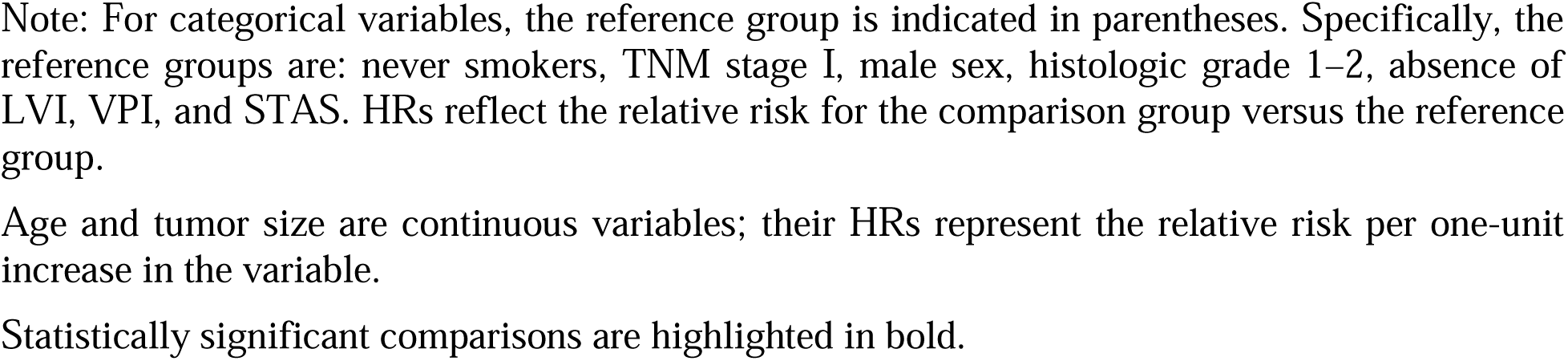
Multivariable Cox regression evaluating overall survival and recurrence-free survival in both the discovery and validation cohorts.

VPI showed a trend toward poorer OS (HR = 1.226, 95% CI = 0.845–1.589, p = 0.075) and RFS (HR = 1.228, 95% CI = 0.912–1.654, p = 0.086), but these associations were not statistically significant.

**Within the validation cohort,** the data were available to calculate both OS and RFS for all 606 patients. The multivariable Cox regression analyses confirmed the independent prognostic value of histologic grade, LVI and STAS for both endpoints **(Table 2)**. Specifically, STAS showed an independent correlation with OS (HR = 1.419, 95% CI = 1.097–1.818, p = 0.029) and with RFS (HR = 1.528, 95% CI = 1.186–1.808, p = 5.38×10□□). LVI also remained significantly associated with OS (HR = 1.140, 95% CI = 0.865–1.510, p = 0.041), but not with RFS. Nevertheless, VPI did not show a significant association with OS or RFS.

These results emphasize the independent prognostic value of grade, LVI and STAS, with grade, and LVI demonstrating the strongest and most consistent associations with survival outcomes, STAS contributing additional independent prognostic information, and VPI showing weaker associations once adjusted for other clinicopathologic features.

### Model Performance and Reproducibility Assessment

The C-index and AUC, which quantify the ability of a model to correctly rank survival times, were used to evaluate the discriminative performance of the different prognostic models **(Tables 3 and S3)**. Prognostic models were first constructed using the 9^th^ edition TNM staging alone, after which histologic factors including histologic grade, STAS, and LVI were sequentially incorporated to assess their incremental contribution to predictive performance.

**Table 3:**
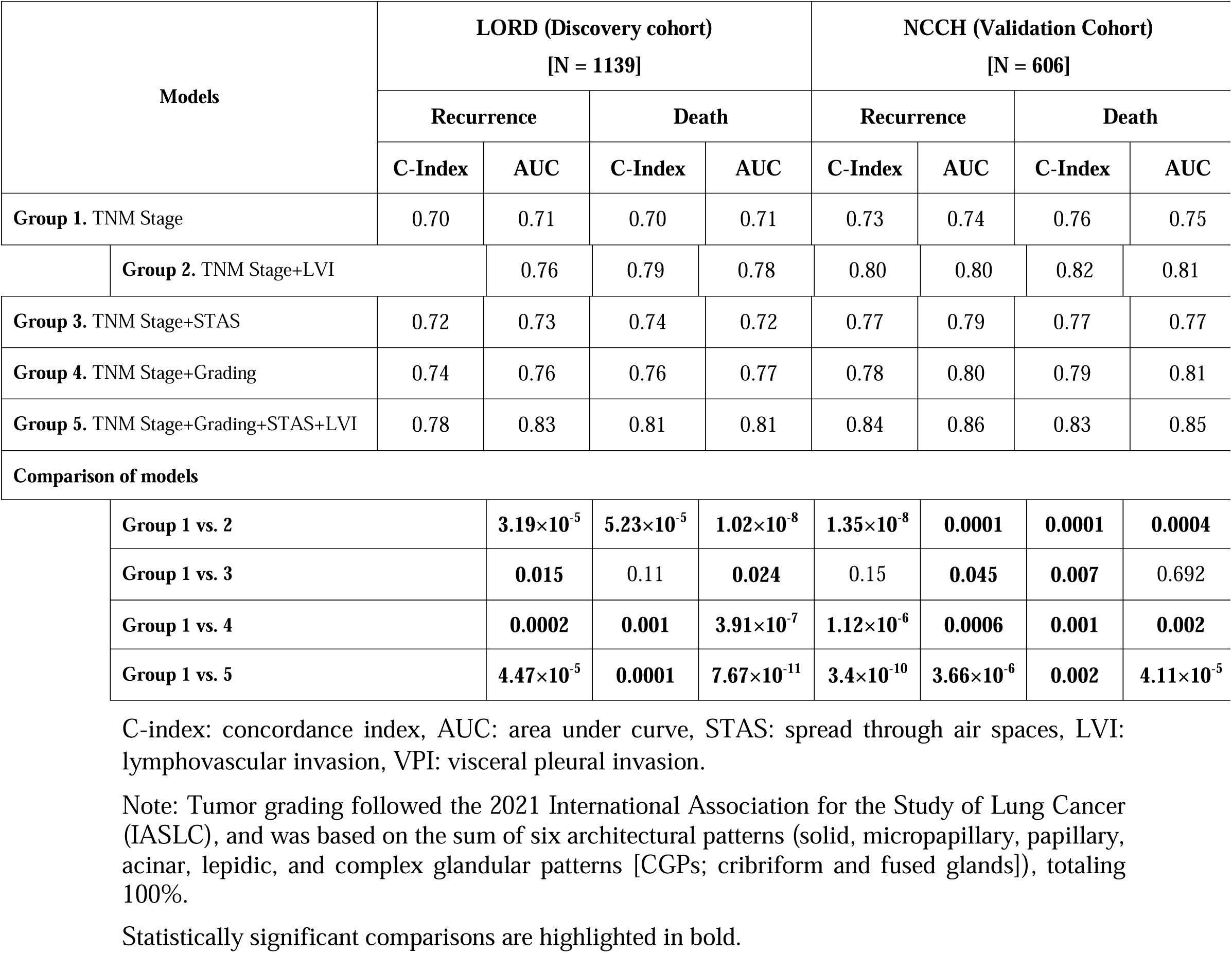
Prognostic model performance based on TNM staging with and without histologic descriptors (Histologic grade, STAS, LVI, VPI).

**In the discovery cohort**, incorporation of LVI into the TNM staging system (Group 2: TNM + LVI) resulted in a significant improvement in prognostic performance compared with TNM alone (Group 1), for both OS (p = 1.02×10^-8^) and RFS (p = 3.19×10^-5^). Addition of STAS alone (Group 3: TNM + STAS) led to a more modest improvement, which reached statistical significance for OS (p = 0.024) and RFS (p = 0.015), but with smaller effect sizes than those observed for LVI.

Inclusion of histologic grade (Group 4: TNM + Grade) further enhanced model discrimination relative to TNM alone, with significant gains for both OS (p = 3.91×10^-7^) and RFS (p = 0.0002). The most comprehensive model, integrating TNM stage, histologic grade, STAS, and LVI (Group 5), demonstrated the strongest overall performance, with significant improvements over TNM alone for both OS (p = 7.67×10^-11^) and RFS (p = 4.47×10^-5^).

**In the validation cohort**, similar trends were observed. Addition of LVI to TNM (Group 2) significantly improved prediction of both OS (p = 0.0004) and RFS (p = 0.0001). Incorporation of histologic grade (Group 4) again resulted in significant improvements for both OS and RFS (OS: p = 0.002; RFS: p = 0.0006). In contrast, STAS alone (Group 3) significantly improved RFS prediction (p = 0.045) but did not improve OS prediction (p = 0.692).

Consistent with the discovery cohort, the combined model including TNM stage, histologic grade, STAS, and LVI (Group 5) demonstrated the highest prognostic accuracy in the validation cohort, with significant improvements over TNM alone for both OS (p = 4.11×10^-5^) and RFS (p = 3.66×10^-6^).

Across both cohorts, the stepwise integration of LVI, histologic grade, and STAS resulted in progressive improvements in C-index and AUC, with LVI and histologic grade contributing the largest gains and STAS providing additional complementary prognostic information. These consistent performance increases across independent cohorts confirm the robustness and reproducibility of the incremental prognostic value conferred by these histologic factors beyond TNM staging alone.

## Discussion

Proposed revisions to the lung cancer TNM classification were developed, and subsequently submitted to the UICC and the AJCC [3, 32, 33]. The integration of histopathologic features into established prognostic frameworks has long been a subject of debate in thoracic oncology. In this study, leveraging a large, multi-sites, and meticulously characterized cohort of NM-LUAD patients treated with complete surgical resection, we comprehensively evaluated the prognostic value of histologic grade, LVI, and STAS alongside VPI, within the framework of the 9^th^ edition TNM staging. Rather than prioritizing a single descriptor, a central aim of our study was to determine the relative and complementary prognostic contributions of histologic grade, LVI, and STAS in combination with established histologic features. This approach aligns with current practices by the AJCC and the UICC, which already recognize certain pathologic features, such as VPI, as prognostically relevant within TNM staging guidelines [3, 11, 34, 35].

### Relative Prognostic Strength of Histologic Descriptors

In our multivariable survival analysis, traditional prognostic factors including TNM stage, tumor size, histologic grade, and LVI demonstrated stronger and more consistent hazard ratios than STAS, while VPI showed the weakest independent associations. Among these, histologic grade, and LVI demonstrated the strongest independent association with poor outcomes for both RFS and OS, aligning with results from multiple studies highlighting its significance in early and advanced-stage LUAD [5, 14, 18, 20, 28, 36]. This hierarchy is expected, as LVI represents well-established marker of biologically aggressive tumor with direct vascular and lymphatic dissemination [5, 8, 11, 14, 18, 28, 36].

### Stage-Dependent Prognostic Effects of histologic grade, LVI, and STAS

Importantly, our stage-stratified analyses demonstrate that the prognostic impact of histologic grade, LVI, and STAS is not uniform across TNM stages, a pattern that has direct implications for their integration into contemporary staging systems. Histologic grade and LVI showed the most robust and consistent prognostic effects, particularly in early-stage disease. High histologic grade was strongly associated with both OS and RFS in stage I tumors, indicating that grading captures aggressive tumor biology even at an early stage. Similarly, LVI demonstrated a consistent association with OS across all stages and with RFS predominantly in stage I disease, supporting its role as a core marker of tumor aggressiveness throughout disease progression.

In contrast, STAS exhibited a more stage-dependent prognostic profile, with a stronger association with adverse outcomes in patients with stage II–III disease and an attenuated, non-significant effect in stage I tumors. This pattern suggests that STAS may reflect invasive tumor behavior that becomes clinically consequential in the context of more advanced disease, rather than functioning as a uniform predictor across all stages. These findings are consistent with large cohort studies and meta-analyses reporting that the prognostic relevance of STAS is variable and influenced by tumor stage, surgical approach, and interaction with other histologic features [37, 38].

Notably, the observed stage-specific effects of histologic grade, LVI, and STAS do not undermine their relevance as potential staging descriptors. Rather than serving as standalone or dominant predictors, histologic grade, LVI, and STAS capture distinct and complementary biological processes that contribute to tumor aggressiveness at different phases of disease evolution.

### Association with Histologic grade

Consistent with prior literature [12, 17, 18, 39–43], we observed that the prevalence of invasive histologic descriptors, including STAS, LVI, and VPI, increased with advancing histologic grade and TNM stage, underscoring their close association with biologically aggressive tumor behavior. This pattern is biologically plausible, as high-grade LUADs are characterized by loss of architectural organization and increased invasive potential, which may facilitate aerogenous, lymphatic, and pleural spread.

Importantly, while the co-occurrence of high-grade histology with STAS and LVI raises the possibility that these descriptors may partially reflect overlapping aspects of tumor aggressiveness, our multivariable analyses demonstrate that both STAS and LVI retain prognostic significance after adjustment for histologic grade. This finding supports the interpretation that invasive histologic features provide complementary prognostic information beyond histologic differentiation alone, rather than serving as redundant surrogates of grade.

The relatively high proportion of grade 3 tumors in the discovery cohort likely reflects multiple factors. Tumor grading was performed according to the 2021 IASLC grading system, in which grade 3 is defined by the presence of ≥20% high-grade patterns (solid, micropapillary, or complex glandular), a definition that may increase the proportion of tumors classified as grade 3 compared with earlier approaches [19–22]. In addition, the discovery cohort spans cases dating back to 2006, when patients more frequently presented with larger and more advanced tumors, whereas contemporary practice increasingly identifies small ground-glass–predominant lesions that are more often low grade [44]. Differences in population characteristics between North American and Asian cohorts, including variation in molecular profiles and associated growth patterns, may also contribute to differences in grade distribution between cohorts.

### Context-Dependent Prognostic Role of STAS and Surgical Approach

The prognostic relevance of STAS has been shown to be highly context-dependent, particularly with respect to surgical approach and extent of resection [37, 38, 45–47]. A recent study by Eguchi et al. [26] confirmed that STAS is associated with a higher risk of recurrence, particularly in patients undergoing sublobar resections, likely due to the anatomic propensity for tumor cell spread within the alveolar parenchyma. In both cohorts, lobectomy was the predominant surgical procedure, performed in over 70% of cases. Biologically, STAS may reflect intrinsic tumor invasiveness and dissemination potential rather than a purely surgical margin–dependent phenomenon, which may explain its persistent prognostic impact across different surgical approaches, including sublobar resection and lobectomy [45–49]. In addition, consistent with the JCOG0802/WJOG4607L trial [50], STAS was associated with poorer RFS in both lobectomy and segmentectomy (sublobar) subgroups, suggesting that its prognostic relevance extends across different surgical resection types.

Nevertheless, we accounted for surgical approach by including it as an adjustment variable in the multivariable models. This is particularly relevant given prior studies indicating that STAS-positive patients who undergo limited resection are at higher risk of recurrence [26, 39]. By controlling for surgical extent, we aimed to isolate the intrinsic prognostic contribution of STAS, independent of treatment-related factors. The persistence of a statistically significant association between STAS and outcomes after adjustment supports its prognostic relevance, while also underscoring that its effect size is more modest and less consistent than that observed for LVI.

### Limited Prognostic Contribution of VPI

In contrast, VPI-which has been a recognized T descriptor, since the first AJCC TNM classification in 1977 [31], demonstrated a less consistent association with survival outcomes than LVI, STAS, and histologic grade. While significant in univariable analyses, VPI did not maintain independent prognostic significance in multivariable models for either OS or RFS. This observation echoes recent findings suggesting that the prognostic role of VPI may be context-dependent and perhaps less pronounced than previously appreciated when considered alongside more sensitive indicators of tumor invasiveness, particularly LVI and, to a lesser extent, STAS [12, 51]. The histopathologic basis for this discrepancy lies in the anatomical limitations of VPI detection, which depends on tumor proximity to the visceral pleura, whereas STAS and LVI can occur independently of tumor location.

### Reproducibility and Generalizability Across Cohorts

To validate and assess the generalizability of our findings, we evaluated a geographically and ethnically independent external cohort. Across both the discovery and validation cohorts, histologic grade and LVI showed the most robust and consistent prognostic effects, particularly in early-stage disease. High histologic grade was strongly associated with both OS and RFS in stage I tumors. Similarly, LVI demonstrated a consistent association with OS across all stages and with RFS predominantly in stage I disease. In contrast, STAS exhibited a more stage-dependent prognostic profile, with a stronger association with adverse outcomes in patients with stage II–III disease and an attenuated, non-significant effect in stage I tumors.

Notably, the prevalence of key invasive histologic descriptors, including STAS, LVI, and VPI, was broadly comparable between the discovery and validation cohorts. In contrast, differences were observed in other clinicopathologic characteristics, such as smoking status and certain demographic variables, reflecting underlying population-, institutional-, or region-specific differences in tumor presentation and clinical practice patterns. Importantly, despite these variations, both cohorts were predominantly composed of early-stage tumors and demonstrated consistent associations between invasive histologic descriptors and clinical outcomes. This is consistent with earlier international collaborative efforts that emphasize the need for standardized reporting of histologic descriptors in lung cancer staging worldwide [3, 11, 41, 52].

Taken together, these findings suggest that while core prognostic patterns were highly consistent across geographically distinct populations, differences in effect magnitude may reflect cohort-specific features, including clinical management practices, surgical selection, and inherent variability in pathological assessment. Importantly, grading system, LVI, and STAS represent robust and reproducible prognostic parameters, and provide complementary, stage-dependent information that may enhance risk stratification and inform refinement of future TNM staging.

### Model Performance and Incremental Prognostic Value

Our study provides important insights into the hierarchical and complementary prognostic utility of histologic descriptors in surgically resected LUAD. Model performance metrics, including C-index and AUC values, demonstrated that the magnitude and consistency of prognostic improvement were greatest when each histologic grade and LVI were incorporated into the TNM staging system, underscoring their central and robust prognostic roles. The addition of STAS alone to TNM staging also resulted in improved model performance, although the magnitude of improvement was consistently weaker than that observed for histologic grade and LVI. Notably, the highest gains in prognostic accuracy were achieved when histologic grade, LVI, and STAS were integrated simultaneously with TNM staging across both cohorts, indicating that these histologic descriptors provide complementary prognostic information beyond established factors such as VPI. The incremental prognostic benefit was particularly pronounced for RFS, suggesting that histologic descriptors may be more informative for predicting early tumor recurrence than long-term survival outcomes. This observation is consistent with population-based data from the TRACERx study [21] and is clinically relevant, as improved identification of patients at high risk of recurrence may inform adjuvant treatment decisions and postoperative surveillance strategies.

### Implications for Future TNM Staging Systems

Our findings raise important considerations for the forthcoming 10^th^ edition of the TNM staging system. Although LVI and Pn are currently listed as optional descriptors by the AJCC and UICC [8, 9], our results provide further support for the integration of LVI and histologic grade as core prognostic histologic features, given their strong, consistent, and stage-spanning associations with both OS and RFS. In contrast, although STAS demonstrated independent prognostic value, its effect was less consistent than that observed for LVI and grading. Moreover, the incorporation of STAS into formal staging frameworks poses unique challenges.

Unlike LVI and VPI, STAS lacks universally standardized diagnostic criteria and quantification methods, with interobserver variability complicating its integration into routine practice [53, 54]. Nevertheless, standardized reporting of STAS as present or absent, as proposed by the IASLC and adopted in CAP synoptic templates, is a pragmatic first step toward harmonization [12, 13, 55].

### Reproducibility and Practical Considerations in Pathologic Assessment

An important consideration for the clinical implementation of invasive histologic descriptors relates to its reproducibility across observers. While formal interobserver variability metrics were not calculated in the present study, the use of standardized morphologic criteria and consensus review among specialized pulmonary pathologists supports the reproducibility of histologic grade, TNM stage, STAS, LVI, and VPI assessment. Importantly, although STAS has historically been associated with concerns regarding interobserver variability, multiple studies have demonstrated that reproducibility is acceptable when stringent morphologic criteria are applied and assessment is performed on adequately sampled permanent sections [4, 12, 14]. Similarly, vascular invasion was assessed using standardized morphologic criteria, although ancillary elastic stains were not systematically performed, which may influence reproducibility and underscores the need for harmonized diagnostic guidelines when considering its incorporation into staging systems [23, 27].

Notably, STAS should not be evaluated on frozen sections, as frozen-section analysis is limited by restricted tissue sampling, sectioning artifacts, and the focal and heterogeneous distribution of tumor cell spread within air spaces, rather than by interobserver disagreement per se [56]. Similar practical considerations apply to other invasive histologic descriptors. LVI, particularly when assessed on well-fixed permanent sections, has demonstrated reasonable interobserver agreement, although distinguishing lymphatic from vascular invasion may require ancillary stains in selected cases [5, 18, 28]. VPI generally shows higher reproducibility owing to well-defined anatomic criteria based on elastic layer involvement [17, 43]. Together, these considerations highlight that, when assessed under standardized conditions, histologic descriptors such as LVI, STAS, and histologic grade represent robust and reproducible prognostic parameters. Importantly, these descriptors provide complementary, stage-dependent information that may enhance risk stratification and inform refinement of future TNM staging frameworks.

### Study Limitations

There are limitations to our study. The retrospective design and reliance on archival pathology slides may introduce selection and reporting biases. Although stringent inclusion criteria and standardized histologic review mitigate these risks, some degree of heterogeneity is inevitable.

Moreover, our analysis was limited to invasive NM-LUAD; due to limited sample availability, we were unable to evaluate other major subtypes of NSCLC. If STAS, and LVI are to be considered for integration into future TNM staging systems, it is essential that their prognostic relevance be validated across the broader spectrum of NSCLC histologies through dedicated studies.

An additional consideration is the extended accrual period of the discovery cohort (2006–2021), during which important advances in lung cancer management occurred, including refinements in surgical techniques, perioperative care, and the introduction of molecularly targeted and immune-based therapies. However, the majority of patients in our cohort had early-stage disease treated with upfront surgery, and receipt of neoadjuvant therapy was an exclusion criterion, thereby limiting baseline treatment heterogeneity. Furthermore, the consistent prognostic effects of histologic grade, STAS and LVI across both OS and RFS, together with external validation in an independent cohort, support the robustness of these associations despite temporal changes in treatment paradigms. Nevertheless, residual confounding related to evolving management strategies remains an inherent limitation of long-term retrospective studies.

### Future Directions

Molecular correlates of these histologic descriptors, which may further elucidate the biological mechanisms underlying tumor aggressiveness, were not explored in the present study, as our analysis was based exclusively on morphologic criteria. Emerging evidence suggests that genomic alterations and chromosomal instability correlate with both and histologic grade, STAS and LVI, highlighting potential biological links between molecular instability and invasive growth patterns [57, 58]. While molecular features represent promising candidates for future refinements of lung cancer classification and risk stratification, their integration into formal TNM staging remains challenging due to issues related to the impact of targeted-therapies, assay standardization, cost, and global accessibility. In contrast, histopathologic parameters such as STAS, LVI, and histologic grade are routinely assessed worldwide and may represent a pragmatic intermediate step toward biologically informed staging systems, while molecular incorporation continues to evolve through prospective, multi-omics studies [57, 58].

## Conclusion

This study demonstrates that histologic grade, LVI, and STAS provide complementary, stage-dependent prognostic information and supports their consideration as integrated histologic modifiers within the TNM staging framework. The integration of these histologic descriptors enhances prognostic discrimination, particularly for recurrence prediction, supporting their consideration in refined staging frameworks. Prospective validation, consensus on diagnostic criteria, and exploration of molecular correlates will be pivotal in advancing these findings toward clinical implementation.

## Data Availability

Ethics Approval and Consent to Participate:

The study was approved by the ethics committee of the Institut Universitaire de Cardiologie et de Pneumologie de Quebec-Laval University, part of the Quebec Respiratory Health Research Network, approval number #2020-3352, 21871. The entire research was performed in accordance with the Declaration of Helsinki. Consent to Participate was waived due to retrospective type of study.

## Acknowledgments

The authors would like to thank all patients included in this study, and the IUCQP-UL Biobank, part of the Quebec Respiratory Health Research Network, for providing the clinical data.

