## Supplementary Material for "Integrating Histologic Descriptors into the Ninth Edition TNM Staging Improves Prognostic Stratification of Lung Adenocarcinoma"

**Supplementary Table 1.** Distribution of STAS, LVI, and VPI According to Clinicopathologic Characteristics (Discovery cohort, N=1139).

| **Patient/Tumor Characteristic** | **STAS** | | ***p* Value**  **(**effect size**)** | **LVI** | | ***p* Value**  **(**effect size**)** | **VPI** | | ***p* Value**  **(**effect size**)** |
| --- | --- | --- | --- | --- | --- | --- | --- | --- | --- |
|  | **Present**  **(N =494)** | **Absent**  **(N =645)** |  | **Present**  **(N =534)** | **Absent**  **(N =605)** |  | **Present**  **(N =311)** | **Absent**  **(N =828)** |  |
| **Sex** | | | | | | | | | |
| Female | 285 (42.6%) | 384 (57.4%) | 0.531  (0.018) | 292 (43.6%) | 377 (56.4%) | **0.009**  (0.077) | 178 (26.6%) | 491 (73.4%) | 0.528  (0.018) |
| Male | 209 (44.5%) | 261 (55.5%) |  | 245 (52.1%) | 228 (48.0%) |  | 133 (28.3%) | 337 (71.7%) |  |
| **Age (y)** | | | | | | | | | |
| ≥ 65 | 379 (43.4%) | 495 (56.6%) | 0.749  (0.02) | 395 (45.2%) | 479 (54.8%) | **0.038**  (0.03) | 227 (26.0%) | 647 (74.0%) | 0.066  (0.04) |
| < 65 | 115 (43.4%) | 150 (56.6%) |  | 139 (52.5%) | 126 (47.5%) |  | 84 (31.7%) | 181 (68.3%) |  |
| **Smoking status** | | | | | | | | | |
| Never | 24 (35.3%) | 44 (64.7%) | 0.165  (0.049) | 20 (29.4%) | 48 (70.6%) | **0.002**  (0.11) | 13 (19.1%) | 55 (80.9%) | 0.118  (0.047) |
| Current/former | 470 (43.9%) | 601 (56.1%) |  | 514 (48.0%) | 557 (52.0%) |  | 298 (27.8%) | 773 (72.2%) |  |
| **Surgical procedure** | | | | | | | | | |
| Lobectomy | 390 (43.8%) | 500 (56.2%) | 0.568  (0.031) | 423 (47.5%) | 467 (52.5%) | **0.00002**  (0.136) | 243 (27.3%) | 647 (72.7%) | **0.021**  (0.082) |
| Segmentectomy | 64 (44.1%) | 81 (55.9%) |  | 47 (32.4%) | 98 (67.6%) |  | 30 (20.7%) | 115 (79.3%) |  |
| Other | 40 (38.5%) | 64 (61.5%) |  | 64 (61.5%) | 40 (38.5%) |  | 38 (36.5%) | 66 (63.5%) |  |
| **Tumor grade** | | | | | | | | | |
| 1 or 2 | 114 (26.9%) | 310 (73.1%) | **<0.00001**  (0.277) | 90 (21.2%) | 334 (78.8%) | **<0.00001**  (0.408) | 63 (14.9%) | 361 (85.1%) | **<0.00001**  (0.231) |
| 3 | 380 (53.2%) | 335 (46.8%) |  | 444 (62.1%) | 271 (37.9%) |  | 248 (34.7%) | 467 (65.3%) |  |
| **Tumor location** | | | | | | | | | |
| Left Upper Lobe | 129 (43.6%) | 167 (56.4%) | 0.991  (0.015) | 145 (49.0%) | 151 (51.0%) | **0.041**  (0.093) | 73 (24.7%) | 223 (75.3%) | 0.581  (0.050) |
| Left Lower Lobe | 60 (41.7%) | 84 (58.3%) |  | 51 (35.4%) | 93 (64.6%) |  | 42 (29.2%) | 102 (70.8%) |  |
| Right Upper Lobe | 186 (43.3%) | 244 (56.7%) |  | 201 (46.7%) | 229 (53.3%) |  | 122 (28.4%) | 308 (71.6%) |  |
| Right Lower Lobe | 79 (44.4%) | 99 (55.6%) |  | 90 (50.6%) | 88 (49.4%) |  | 45 (25.3%) | 133 (74.7%) |  |
| Other | 40 (44.0%) | 51 (56.0%) |  | 47 (51.6%) | 44 (48.4%) |  | 29 (31.9%) | 62 (68.1%) |  |
| **Categorical tumor size** | | | | | | | | | |
| > 3 cm | 144 (40.1%) | 215 (59.9%) | **0.00004**  (0.044) | 213 (59.3%) | 146 (40.7%) | **0.0001**  (0.169) | 139 (38.7%) | 220 (61.3%) | **<0.00001**  (0.173) |
| ≤ 3 cm | 350 (44.9%) | 430 (55.1%) |  | 321 (41.2%) | 459 (58.8%) |  | 172 (22.1%) | 608 (77.9%) |  |
| **Tumor size** | | | | | | | | | |
| Mean (SD) | 2.69 (1.52) | 2.81 (1.85) | 0.597  (0.02) | 3.06 (1.9) | 2.5 (1.48) | **<0.00001**  (0.23) | 3.3 (2.15) | 2.56 (1.47) | **<0.00001**  (0.26) |
| Median (Range) | 2.3 (1-12.5) | 2.3 (1-24.5) |  | 2.6 (1-24.5) | 2.2 (1.12.5) |  | 2.8 (1-24.5) | 2.2 (1-15.0) |  |
| **TNM stage** | | | | | | | | | |
| I | 346 (41.7%) | 483 (58.3%) | 0.137  (0.058) | 312 (37.6%) | 517 (62.4%) | **<0.00001**  (0.303) | 178 (21.5%) | 651 (78.5%) | **<0.00001**  (0.246) |
| II | 96 (46.2%) | 112 (53.8%) |  | 149 (71.6%) | 59 (28.4%) |  | 74 (35.6%) | 134 (64.4%) |  |
| III | 52 (51.0%) | 50 (49.0%) |  | 73 (71.6%) | 29 (28.4%) |  | 59 (57.8%) | 43 (42.2%) |  |
| **STAS** | | | | | | | | | |
| Negative | ----- | | | 255 (39.5%) | 390 (60.5%) | **<0.00001**  (0.168) | 175 (27.1%) | 470 (72.9%) | 0.881  (0.004) |
| positive |  |  |  | 279 (56.5%) | 215 (43.5%) |  | 136 (27.5%) | 358 (72.5%) |  |
| **LVI** | | | | | | | | | |
| No invasion |  | |  | ----- | | | 91 (15.0%) | 514 (85.0%) | **<0.00001**  (0.292) |
| Invasion | ----- | |  |  |  |  | 220 (41.2%) | 314 (58.8%) |  |

LVI: lymphovascular invasion, STAS: spread through air spaces, VPI: visceral pleural invasion.

Note: Cohort includes all patients undergoing R0 resection for LUAD with known STAS, LVI, and VPI status.

Bold emphasis is used to indicate statistically significant comparisons.

The p-value were calculated with Mann-Whitney U test (Wilcoxon rank-sum test) for age and tumor size, and Chi-square test or Fisher’s exact test for the other variables.

The effect sizes in the parentheses below the p-value, were calculated with Rank-biserial correlation for age and tumor size, and the Cramer’s V (for larger tables) or Phi coefficient (for 2×2 tables) for categorical variables.

**Supplementary Table 2:** Proportion With STAS, LVI, and VPI by Stage and LUAD Histological Grade (IUCPQ cohort, N=1139)

| **LUAD grade** | **N** | **STAS** | | *p* Value | **LVI** | | *p* Value | **VPI** | | *p* Value |
| --- | --- | --- | --- | --- | --- | --- | --- | --- | --- | --- |
|  |  | **Absent** | **Present (%)** |  | **Absent** | **Present (%)** |  | **Absent** | **Present (%)** |  |
| **Stage I**  Grade 1  Grade 2  Grade 3 | 103  271  455 | 91  185  207 | 12 (11.65%)  86 (31.73%)  248 (54.51%) | **<0.0001** | 97  211  209 | 6 (5.83%)  60 (22.14%)  246 (54.07%) | **<0.0001** | 99  230  322 | 4 (3.88%)  41 (15.13%)  133 (29.23%) | **<0.0001** |
| **Stage II**  Grade 1  Grade2  Grade 3 | 1  36  171 | 1  26  85 | 0 (0%)  10 (27.78%)  86 (50.29%) | **<0.0001** | 0  18  41 | 1 (100%)  18 (50%)  130 (76.02%) | **<0.0001** | 1  22  111 | 0 (0%)  14 (38.89%)  60 (35.09%) | **<0.0001** |
| **Stage III**  Grade 1  Grade 2  Grade 3 | 0  13  89 | 0  7  43 | 0 (0%)  6 (46.15%)  46 (51.69%) | **<0.0001** | 0  8  21 | 0 (0%)  5 (38.46%)  68 (76.4%) | **<0.0001** | 0  9  34 | 0 (0%)  4 (30.77%)  55 (61.8%) | **<0.0001** |

LVI: lymphovascular invasion, STAS: spread through air spaces, VPI: visceral pleural invasion.

Note: The p-value were calculated with Chi-square test or Fisher’s exact test.

Bold emphasis is used to indicate statistically significant comparisons.

In Stage I, the prevalence of STAS, LVI, and VPI increased progressively with tumor grade, observed in 11.7%, 31.7%, and 54.5% for STAS; 5.8%, 22.1%, and 54.1% for LVI; and 3.9%, 15.1%, and 29.2% for VPI, respectively, from grades 1 to 3 (all p < 0.0001). A similar trend was noted in Stage II, where STAS was absent in grade 1 but detected in 27.8% and 50.3% of grade 2 and 3 tumors, respectively; LVI was likewise absent in grade 1 and observed in 50.0% and 76.0% of grades 2 and 3; while VPI was not observed in grade 1, and present in 38.9% and 35.1% of grades 2 and 3 tumors (all p < 0.0001). In Stage III, none of the grade 1 tumors exhibited STAS, LVI, or VPI, whereas STAS was present in 46.2% and 51.7% of grades 2 and 3; LVI in 38.5% and 76.4%; and VPI in 30.8% and 61.8%, respectively (all p < 0.0001).

**Supplementary Table 3:** Prognostic model performance based on TNM staging with and without histologic descriptors (Tumor grade, STAS, LVI, VPI).

| **Model** | **IUCPQ (Discovery cohort)**  **[N = 1139]** | | | | **NCCH (Validation Cohort)**  **[N = 606]** | | | |
| --- | --- | --- | --- | --- | --- | --- | --- | --- |
|  | **Recurrence** | | **Death** | | **Recurrence** | | **Death** | |
|  | **C-index** | **AUC** | **C-index** | **AUC** | **C-index** | **AUC** | **C-index** | **AUC** |
| **Tumor Grade** | 0.69 | 0.70 | 0.71 | 0.72 | 0.72 | 0.74 | 0.74 | 0.75 |
| **STAS** | 0.65 | 0.66 | 0.66 | 0.67 | 0.69 | 0.70 | 0.70 | 0.71 |
| **LVI** | 0.71 | 0.73 | 0.73 | 0.74 | 0.75 | 0.77 | 0.76 | 0.78 |
| **TNM stage** | 0.70 | 0.71 | 0.70 | 0.71 | 0.73 | 0.74 | 0.76 | 0.75 |
| **TNM stage+ Grade** | 0.74 | 0.76 | 0.76 | 0.77 | 0.78 | 0.80 | 0.79 | 0.81 |
| **TNM stage+ STAS** | 0.72 | 0.73 | 0.74 | 0.72 | 0.77 | 0.79 | 0.77 | 0.77 |
| **TNM stage+ LVI** | 0.76 | 0.79 | 0.78 | 0.80 | 0.80 | 0.82 | 0.81 | 0.83 |
| **TNM stage+ STAS + LVI** | 0.77 | 0.81 | 0.80 | 0.81 | 0.82 | 0.85 | 0.82 | 0.83 |
| **TNM stage+ Grade + STAS** | 0.77 | 0.82 | 0.80 | 0.82 | 0.83 | 0.85 | 0.82 | 0.84 |
| **TNM stage+ Grade + LVI** | 0.78 | 0.82 | 0.81 | 0.83 | 0.84 | 0.86 | 0.83 | 0.85 |
| **TNM stage+ Grade + STAS + LVI** | 0.78 | 0.83 | 0.81 | 0.81 | 0.84 | 0.86 | 0.83 | 0.85 |

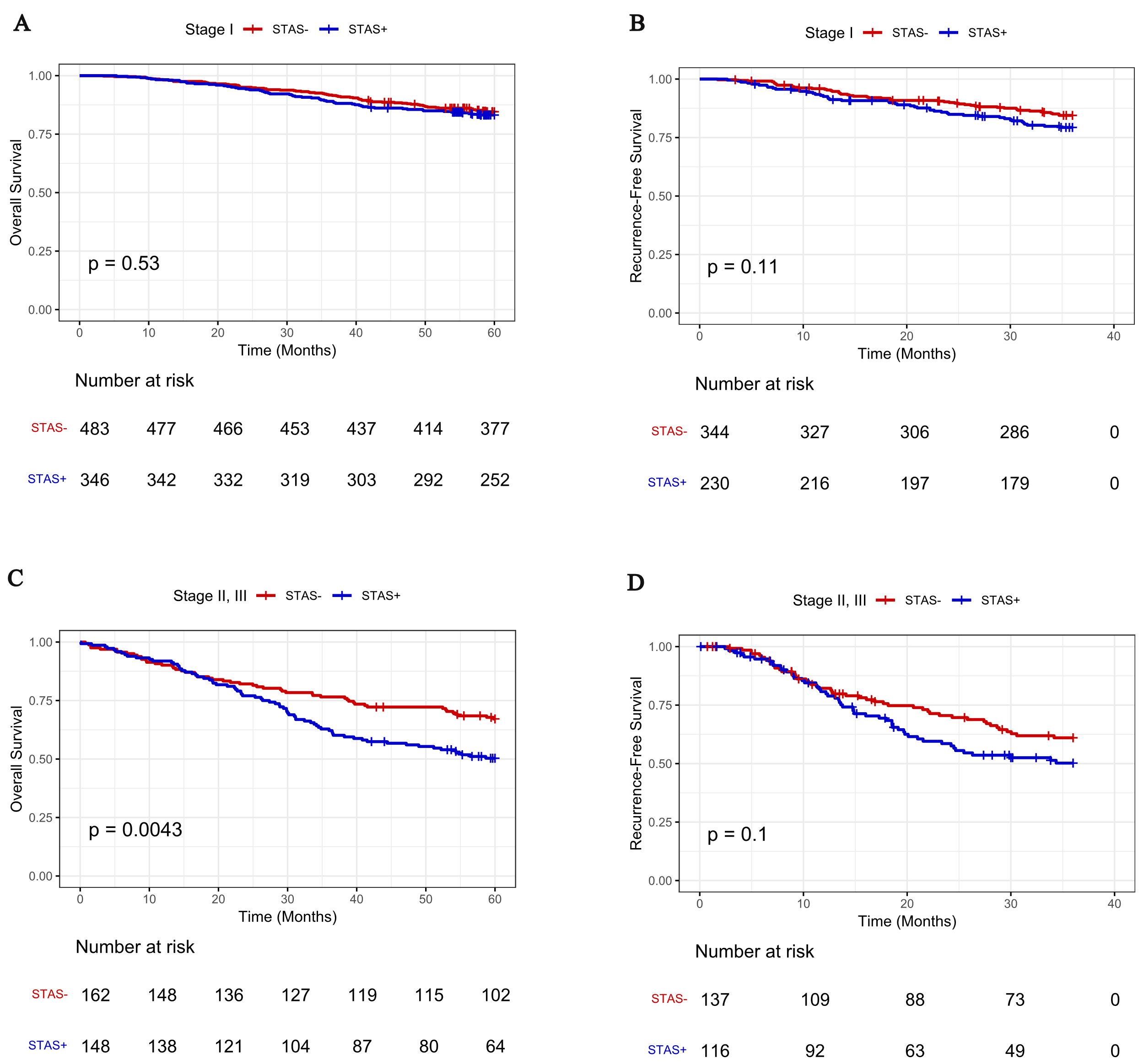

**Supplementary Figure 1: (A, B)** Prognostic Impact of STAS on overall survival (OS), and recurrence-free survival (RFS) in stage I LUAD patients. **(C, D)** Prognostic Impact of STAS on OS, and RFS in stage II, and III LUAD patients. STAS: spread through air spaces.

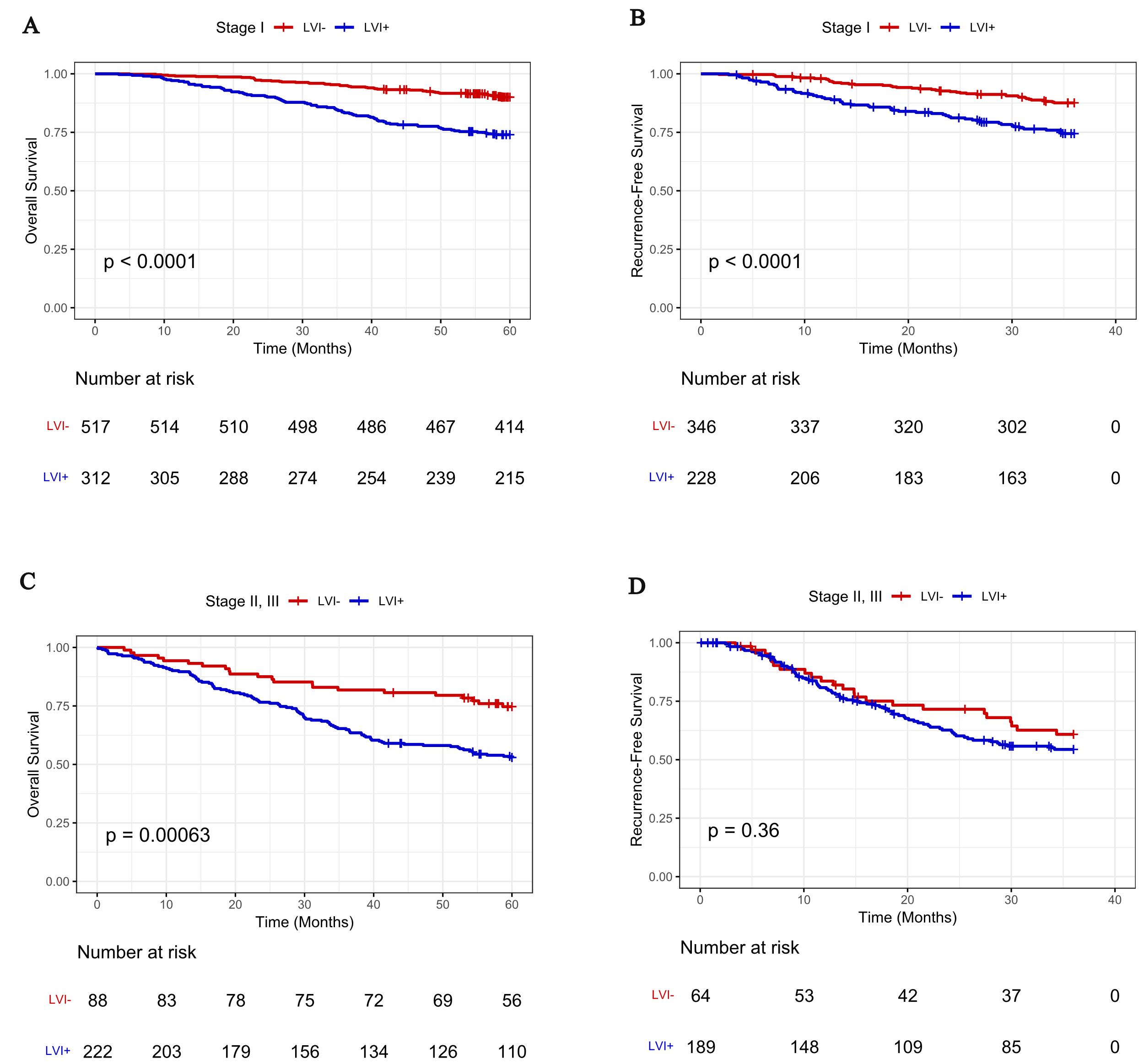

**Supplementary Figure 2: (A, B)** Prognostic Impact of LVI on overall survival (OS), and recurrence-free survival (RFS) in stage I LUAD patients. **(C, D)** Prognostic Impact of LVI on OS, and RFS in stage II, and III LUAD patients. LVI: lymphovascular invasion.

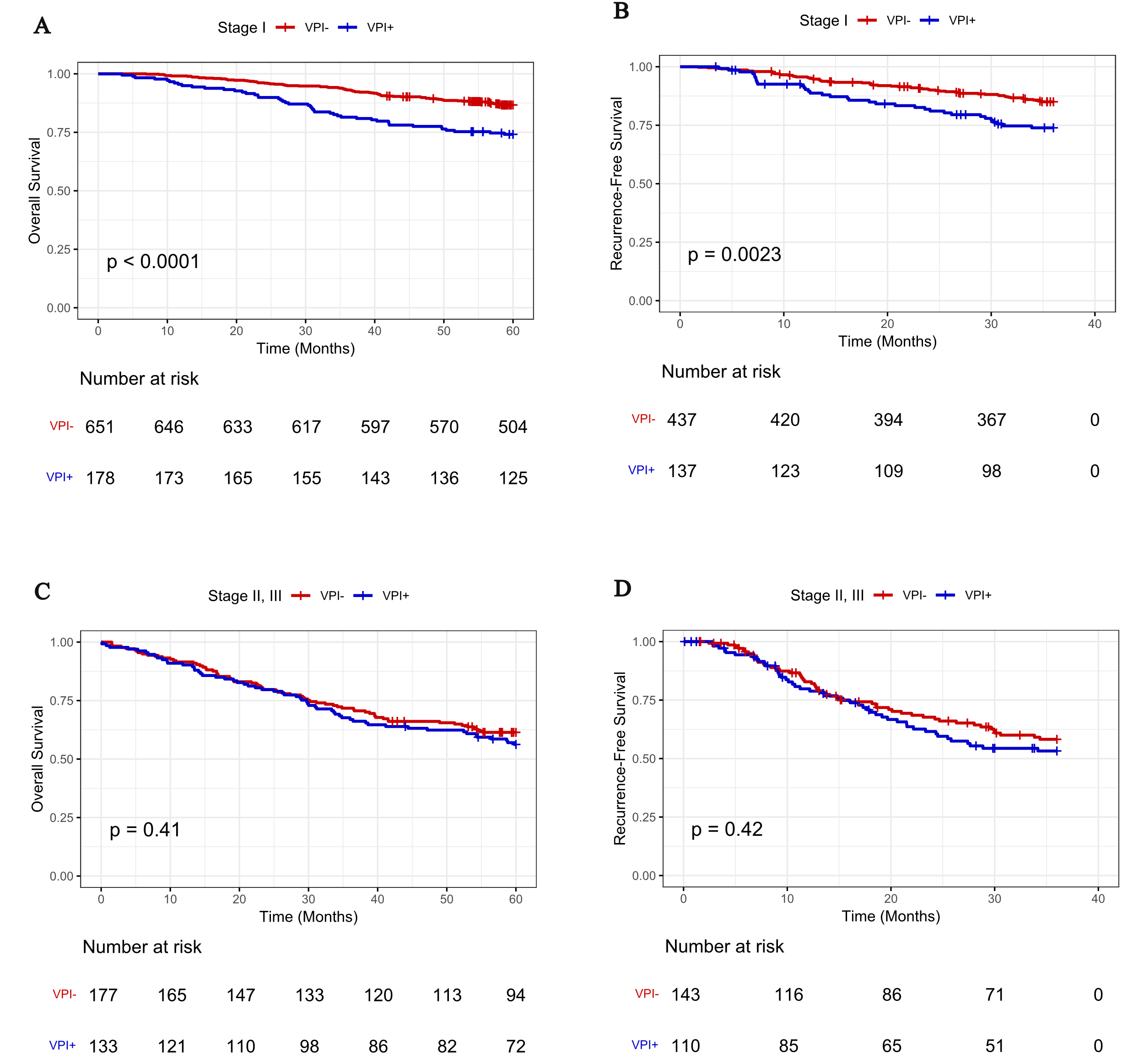

**Supplementary Figure 3: (A, B)** Prognostic Impact of VPI on overall survival (OS), and recurrence-free survival (RFS) in stage I LUAD patients. **(C, D)** Prognostic Impact of VPI on OS, and RFS in stage II, and III LUAD patients. VPI: visceral pleural invasion.

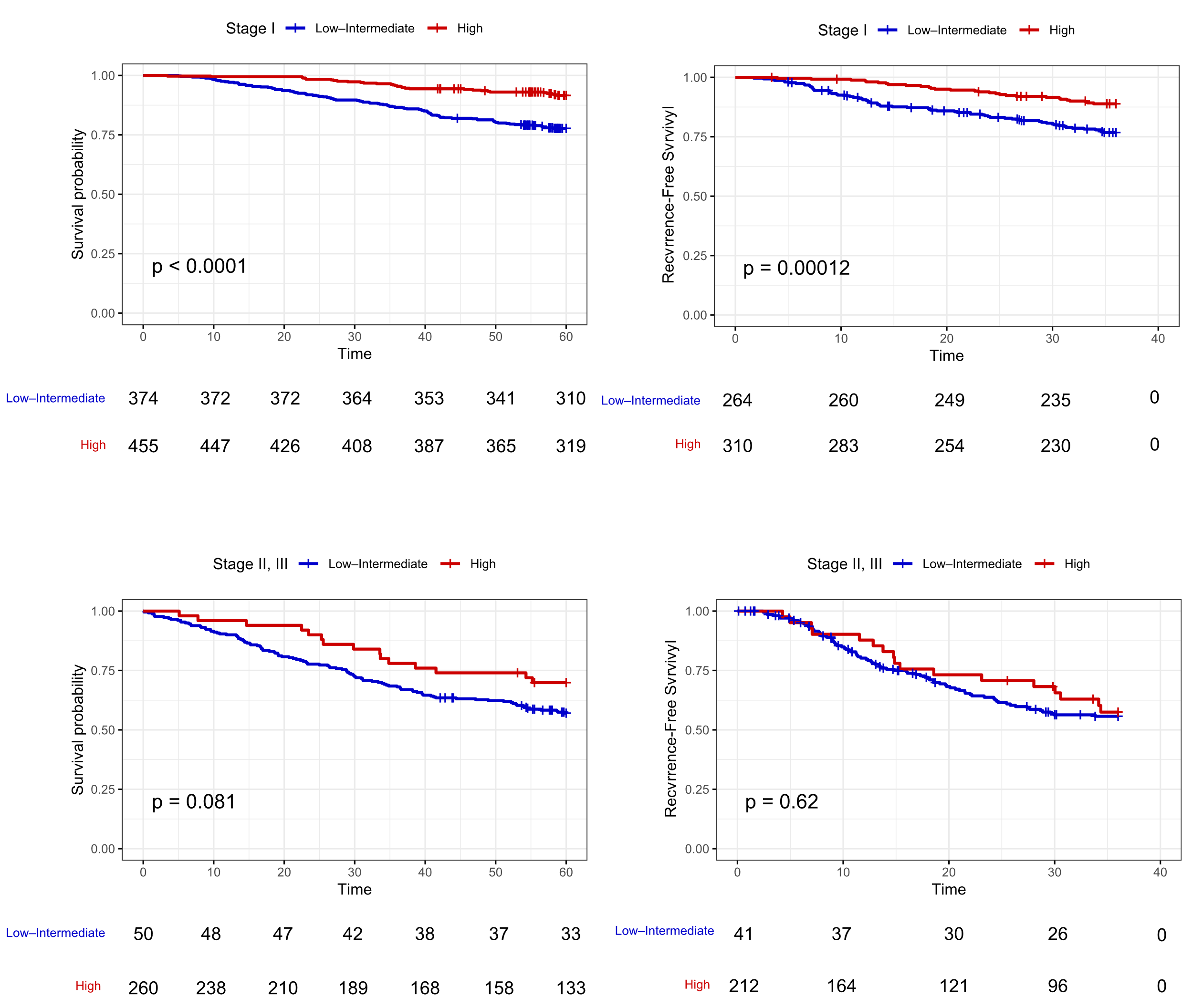

**Supplementary Figure 4: (A, B)** Prognostic Impact of tumor grade on overall survival (OS), and recurrence-free survival (RFS) in stage I LUAD patients. **(C, D)** Prognostic Impact of tumor grade on OS, and RFS in stage II, and III LUAD patients.
